# Tooth loss, diet and cardiovascular disease: A longitudinal study in middle-aged Australian women

**DOI:** 10.64898/2025.12.14.25342227

**Authors:** Shalinie King, Simone Marschner, Desi Quintans, Alice Gibson, Clara K Chow

**Author notes:** Corresponding author: The University of Sydney, Westmead Hospital, 176 Hawkesbury Road, Westmead 2145, Sydney, Australia., **Email:**.

## Abstract

**Background and objective:** Cardiovascular disease (CVD) is highly prevalent affecting one in six Australians. Tooth loss has consistently been associated with an increased risk of CVD, influenced by social determinants of health as well as effects on chewing function and diet. However, there is little evidence regarding the role of diet on the relationship between tooth loss and CVD. This study aimed to determine the impact of retaining a functional dentition (minimum of 20 teeth) on the risk of incident CVD and the role of diet in this relationship.

**Methods:** This prospective cohort study recruited Australian women from the 1946-51 cohort of the Australian Longitudinal Study on Women’s Health. Tooth loss and diet quality were assessed from survey 5 (2007) and incident CVD from linked data over a 17-year follow-up period (2024). Cox proportional hazards models were used to estimate hazard ratios (HRs) and 95% confidence intervals (CIs) adjusted for known risk factors (age, sociodemographic, lifestyle and medical history factors).

**Results:** A total of 8306 women, average age 58.5 ± 1.5 years, 69.2% with a functional dentition were included. Over the 17-year follow-up period 1432 (17.2%) women developed incident CVD. The presence of a functional dentition was associated with a lower risk of incident CVD compared to women with a non-functional dentition, 15.5% vs 21.1% respectively, [HR 0.83 (95% CI 0.74, 0.93)]. A greater proportion of women with a functional dentition had a high-quality diet score, compared to those with a non-functional dentition (42% vs 36%, p <0.001). Controlling for diet quality did not attenuate this association [HR 0.82 (95% CI 0.73, 0.92)].

**Conclusion:** Although tooth loss negatively impacts diet quality, dietary changes alone do not explain why tooth loss is a marker of incident CVD and loss of a functional dentition should prompt cardiovascular prevention action.

**What is already known on this topic:**

- Previous studies have shown consistent associations between tooth loss and both a higher risk of cardiovascular disease, and poor diet quality. The role of diet in the relationship between tooth loss and cardiovascular disease is unclear.

**What this study adds:**

- This prospective analysis of 8306 Australian women shows that maintaining a functional dentition (minimum 20 teeth) is associated with a lower risk of incident cardiovascular disease.
- Retaining teeth as part of a functional dentition is associated with a better diet quality.
- The relationship between tooth loss and cardiovascular disease is not attenuated when adjusted for diet.

**How this study might affect research, practice or policy:**

- Loss of a functional dentition should serve as a trigger for cardiovascular preventive measures
- Dietary changes alone do not explain why tooth loss is a marker of incident cardiovascular disease, further research on the role of systemic inflammation is required.

## Introduction

Cardiovascular disease (CVD) is a prevalent health condition, representing 32% of all deaths globally.[1] In Australia one in six people self-report that they live with CVD representing almost 18% of the total Australian population.[2]

Oral diseases are also highly prevalent affecting nearly 3.7 billion people globally.[3] Untreated, oral diseases such as tooth decay and periodontitis can lead to the loss of teeth. In Australia the proportion of adults with significant tooth loss increases with age from 0.7% in 15-34 year olds to 46% in those aged 75 years or over.[4] Longitudinal studies conducted globally have demonstrated a consistent association between tooth loss and CVD even after adjusting for sociodemographic factors and chronic conditions such as diabetes, and hypertension. These studies have shown that tooth loss is associated with an increased risk of CVD including CVD disease related mortality [5–8] with a dose-dependent relationship between tooth loss and CVD.[7, 9] However very few studies have considered the role of diet in this association.

The mechanisms by which tooth loss impacts the risk of CVD relate to inflammation [10] and compromised chewing function.[11] Periodontitis a chronic inflammatory condition of the supporting tissues of the teeth is triggered by oral bacteria.[12] In addition to localised oral inflammation, periodontitis results in systemic inflammation and is associated with elevated levels of circulating inflammatory cytokines such as c-reactive protein [10] thereby increasing the cardiovascular risk profile.[13] In terms of chewing function, as teeth are lost and the dentition becomes non-functional and individuals alter their food choices choosing softer foods that are often low in protein and fibre.[14] A functional dentition required to maintain satisfactory chewing performance is defined as the presence of a minimum of 20 natural teeth [4] (A complete dentition consists of 28 -32 teeth).

Poor diet quality contributes to an increased risk of CVD related mortality,[15] whilst an increased intake of both protein and fibre has been reported to reduce the risk of CVD.[16, 17] As such, the loss of a functional dentition may compromise diet quality and thereby increase the risk of CVD. Therefore, an important question is whether the association between tooth loss and CVD is impacted by diet quality.

The primary aim of this study was to determine the association between retaining a functional dentition and the risk of incident CVD in a prospective cohort study of Australian women and investigate the role of diet in this relationship. Secondary aims were to investigate the impact of dentition status on diet quality and the association between diet quality and incident CVD.

## Methods

### Study population

This prospective cohort study used data from the Australian Longitudinal Study on Women’s Health (ALSWH), collected from over 40,000 women in three age groups randomly selected in 1996 from the Medicare database [18]. For this study, data from the 1946-51 cohort of women who answered survey 5 in 2007 when they were aged 56-61 years (n= 10638), was used as our initial sample as it was the first time a question on tooth loss was included in the surveys. Demographic data not available in survey 5 (2007), including income and education level were obtained from survey 3 and 1 respectively. The ALSWH survey program has ongoing ethical approval from the Human Research Ethics Committees (HRECs) of the Universities of Newcastle and Queensland (approval numbers H076-0795, 2004/HE000224, respectively) for the 1946-51 cohort. The ALSWH also maintains institutional HREC approvals for external record linkage. The study conforms to the STROBE guidelines.

### Primary endpoint

The primary endpoint was incident CVD assessed using data obtained from the Common Conditions from Multiple Sources (CCMS) dataset which is derived from surveys, and linked with external health record collections including the National Death Index, Medicare Benefits Schedule, Pharmaceutical Benefits Scheme, hospital admissions and National Aged Care data.[18] CVD included ischaemic heart disease (IHD) and stroke as defined by the 10^th^ revision of the International Statistical Classification of Diseases and Related Health Problems (ICD-10). The codes for IHD included ICD-codes 120-125, and 410-414, and for stroke ICD-codes 430-438, and 160-169. Fatal CVD was defined where the death was caused by IHD or stroke. Incident CVD was defined as the first occurrence of either a diagnosis of CVD (from survey data) or a cardiovascular event (from linked data) during the 17-year follow-up period. This follow-up commenced from each participant’s date of submission of Survey 5 and continued until April 2024, excluding individuals with pre-existing CVD identified through Survey 5 or CCMS data linkage prior to that time. Participants who opted out from the CCMS data linkage were excluded from the analysis.

### Primary independent variable

The primary independent variable was the self-reported number of teeth, assessed using data from survey 5. Dentition status was categorised as functional (≥ 20 teeth and at least 10 teeth in each arch) and non-functional (< 20 teeth, or ≥20 teeth with < 10 teeth in one arch). The remaining dentition was further segmented as “≥20,” “10–19,” “1–9’ or “no remaining teeth” to understand the dose response relationship. Participants with missing tooth information were excluded from the analysis.

### Diet quality

Diet quality was measured using a shortened version of the Dietary Questionnaire for Epidemiological Studies version 2 (DQES) [19] which was administered to all participants in survey 5. The original DQES v2 had a 10-point frequency scale which participants used to report their habitual consumption of 74 foods and beverages over the previous 12-month period. Details of the DQES have previously been presented [19] and the survey has been validated [20]. The shortened version uses a 3 -point frequency scale for most questions except for questions on dairy, meat and fish which use a 5-point frequency scale.

A diet score was determined using the Australian Recommended Food score (ARFS) [21] which is calculated by summing points within the subscales for a total of 74 points [Vegetables (/22), Fruits (/14), Protein foods (/14), Grains (/14), Dairy products (/7), Fats (/1) and Alcohol (/2)]. Foods are given 1 point for a frequency of more than once/week. Higher values correspond to a healthier diet quality. The AFRS was categorised into quintiles based on the distribution of the scores. Quintiles 1-3 were categorised as “low quality” and quintiles 4-5 as “high quality”. Missing diet values on the survey were recoded to zero for up to four items. Participants with missing values for greater than four items were considered to have incomplete data and excluded from the analysis.

### Covariates

Covariates were obtained from survey 5 and included age, area of residence (defined as urban cities, inner regional, outer regional, remote, very remote), socio-economic status and education status [defined by the socio economic indexes for areas (SEIFA) index of socioeconomic disadvantage and the SEIFA index of education, respectively; both of which were defined as either high (above the median cut point) or low (below the median cut point)]. Health related variables included BMI (continuous variable), smoking (defined as never, ex-smoker and current smoker), diabetes, and hypertension both of which were categorical.

### Statistical analysis

All statistical analyses were performed using R software (version 4.4.0)[22] and p-values of less than 0.05 were considered statistically significant unless stated otherwise. Continuous variables are presented as mean ± SD and categorical variables as numbers and percentages. Data excluded and missing are described and we compared baseline characteristics in women with and without data on teeth and/or diet information to assess the potential for selection bias.

Kaplan-Meier survival curve was constructed to examine the association between dentition status and incident CVD (with no teeth used as the reference). Time-to-event data were defined as the duration from survey 5 to the first occurrence of CVD. Hazard ratios were calculated from Cox proportional hazard models and data adjusted for all prespecified covariates.

The primary analysis was with Cox proportional-hazards models to calculate the hazard ratios (HRs) and their 95% confidence intervals (CIs). Model 1 was unadjusted, model 2 was adjusted for all pre-specified covariates and model 3 was further adjusted for diet quality.

To investigate the relationship between dentition status and diet quality and to explore the influence of denture use (based on the response to the questions “do you wear an upper/lower denture?”) on the relationship between dentition status and diet, log binomial analysis was used to calculate risk ratios (RR) and their 95% confidence intervals (CIs). Models were adjusted for all covariates. Further analyses examined the relationship between diet quality as a continuous variable or defined by fruit and vegetable intake only and incident CVD using proportional hazards models to calculate HR and their 95% CIs.

## Patient and public involvement

Patients were not involved in the research process of this study.

## Results

After exclusions the final sample included 8306 participants (Figure 1) with an average age of 58.5 ± 1.5 years. Women with a non-functional dentition had an average of 10.7 ± 7.6 teeth, whilst women with a functional dentition had an average of 26.8 ± 2.9 teeth. Compared to women with a functional dentition, women with a non-functional dentition were more likely to reside in a regional area (66.9% vs 54.1%), and less likely to be in the top two quintiles for education status (37.9% vs 57.3%), and socioeconomic disadvantage (39.6% vs 56.3%). Health-wise, those with a non-functional dentition had a higher mean BMI (28.4 ± 5.9 vs 26.7 ± 5.1) and were more likely to have diabetes and hypertension (8.1% vs 4.2% and 30.2% vs 24.0%, respectively). The majority of those with a non-functional dentition had their teeth replaced with dentures (86.4%). Compared to women with a non-functional dentition, women with a functional dentition were more likely to have a diet score in the top two quintiles (42.0% vs 35.6%), indicating a higher diet quality, Table 1, (online supplemental table 1). Missingness for all covariates was less than 2%. Diet variables were normally distributed, data for the number of teeth were not normally distributed.

**Figure 1:**
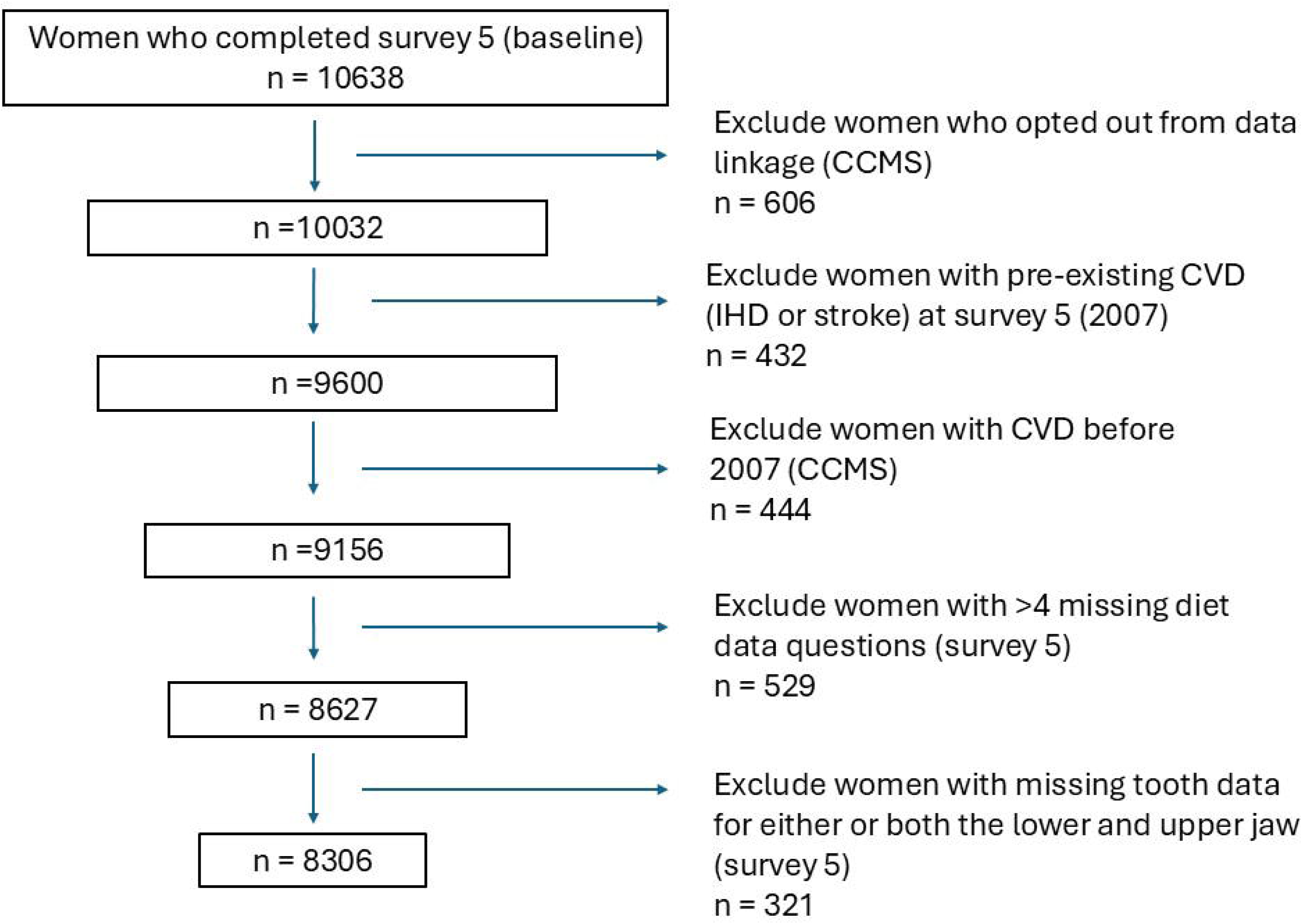
Participant selection flow diagram. Common conditions from multiple sources (CCMS), cardiovascular disease (CVD), ischaemic heart disease (IHD).

**Table 1:**
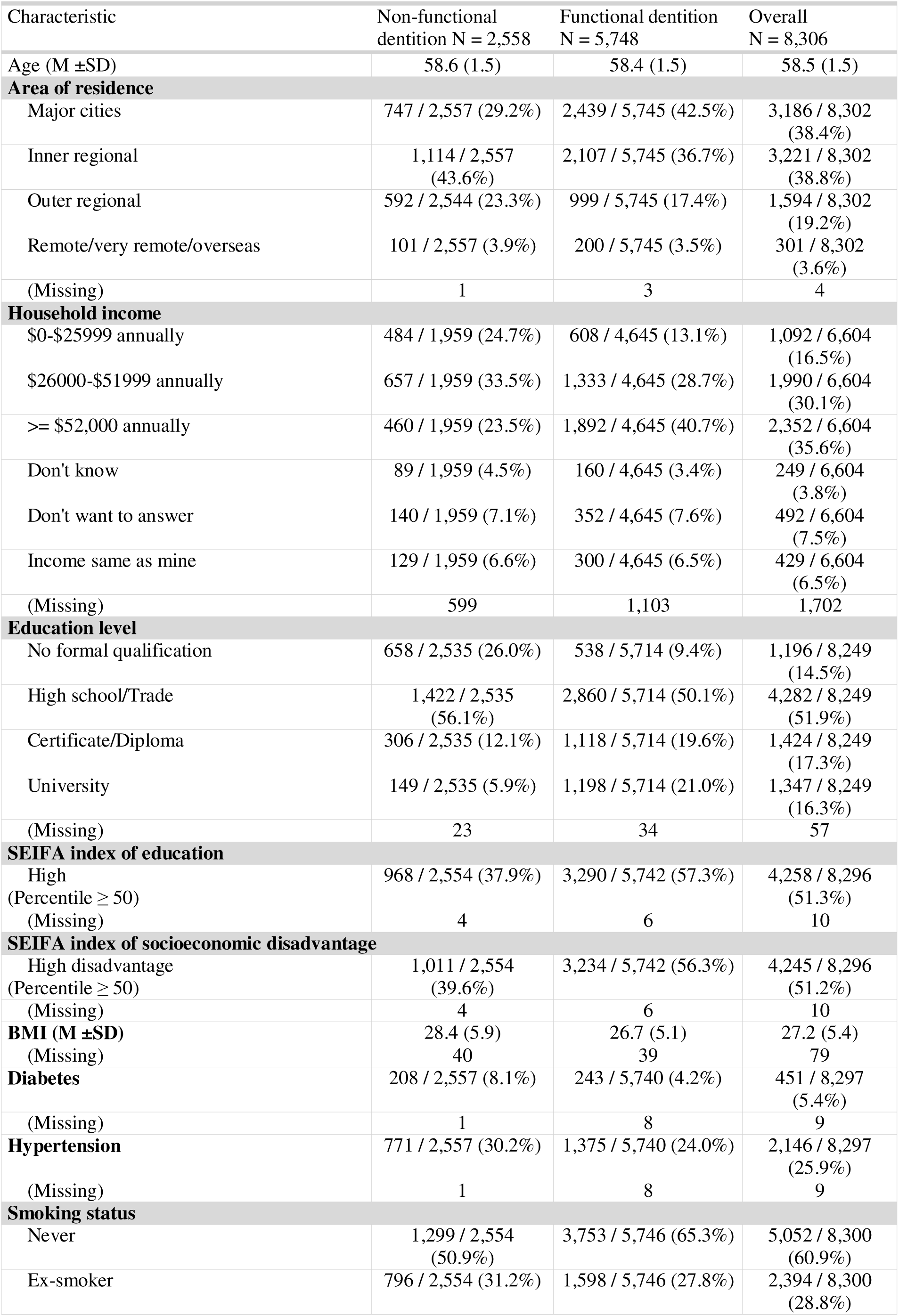
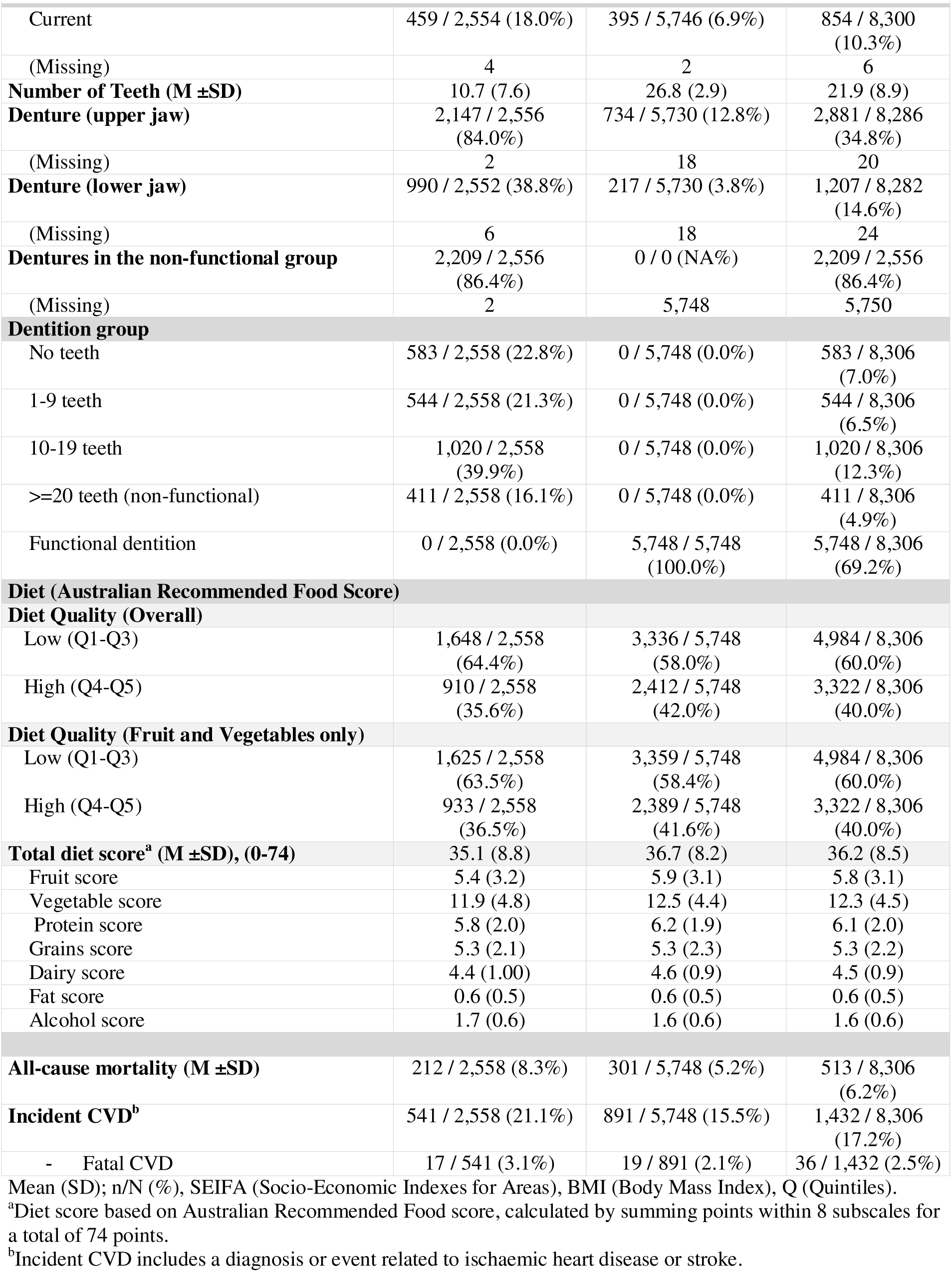
Population Characteristics.

Compared to the study population analysed, women with missing diet and tooth data were older (58.6 ± 1.5 vs 58.5 ± 1.5, p = 0.001), were more likely to live in a rural or remote location (70.3 vs 61.6, p <0.001), more likely to have a low income (21.2% vs 16.5%, p <0.001), and have no formal educational qualification (26.6% vs 14.5%, p<0.001). However, there were no differences in the prevalence of diabetes (p=0.07), hypertension (p=0.12), incident CVD or fatal CVD (p=0.2), (online supplemental table 2).

Over the 17-year follow-up period 513 (6.2%) women died from all causes, 36 of whom due to fatal CVD. A total of 1432 (17.2%) women were diagnosed with CVD (IHD or stroke) or had a cardiovascular event (including fatal CVD).

Women with a non-functional dentition developed CVD earlier and at the 17-year endpoint had a higher rate of incident CVD (Figure 2). A functional dentition was associated with a lower risk of incident CVD in the adjusted model [HR 0.83 (0.74,0.93), p=0.001]. Adjusting for diet quality (high or low) did not attenuate this association [HR 0.82 (95% CI 0.73,0.92), p<0.001], Table 2, (online supplemental table 3). The relationship between the number of teeth and incident CVD suggested a dose response relationship with the HRs for incident CVD decreasing as the number of retained teeth increased (Figure 3).

**Figure 2:**
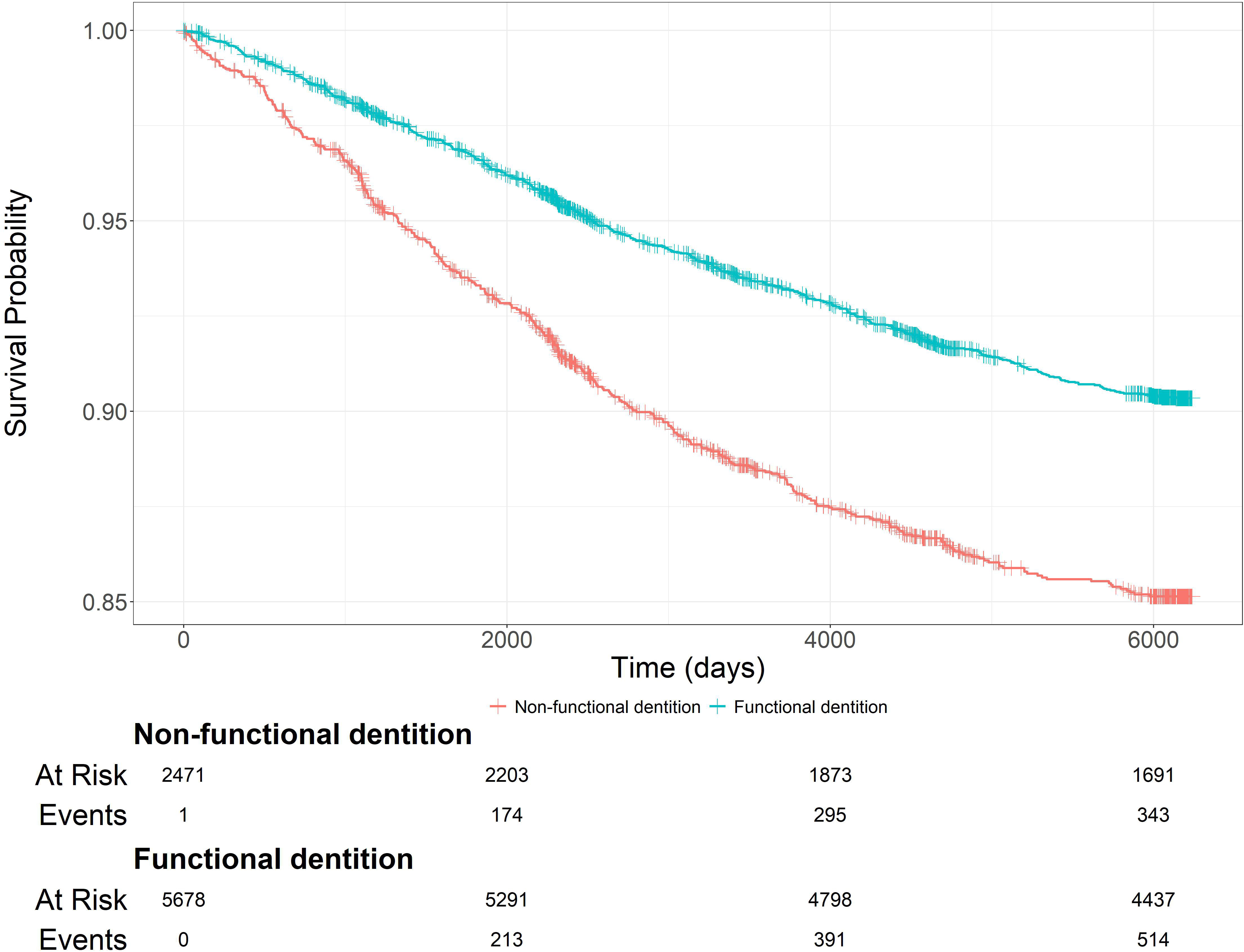
Kaplan-Meier Curve of incident cardiovascular disease across time (in days). The number of participants at risk of incident cardiovascular disease over the 17-year follow up period is shown in women with a functional dentition compared to women with a non-functional dentition.

**Figure 3:**
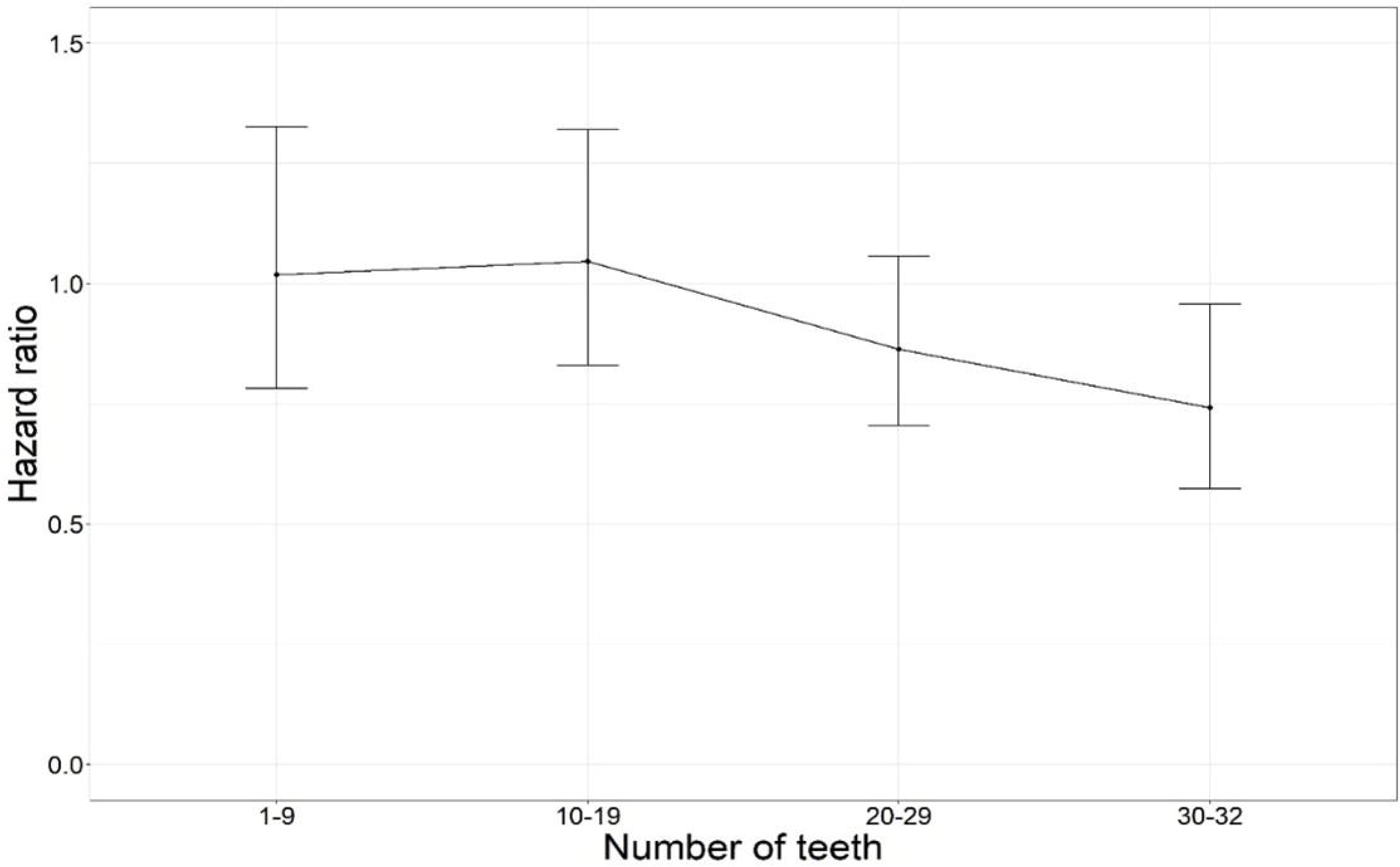
Association between the number of teeth and risk of incident cardiovascular disease. The reference group are women with no teeth. Hazard ratios are calculated from Cox proportional hazard models and data adjusted for adjusted for age, area of residence, SEIFA index of socio-economic disadvantage, SEIFA index of education, body mass index, smoking status, diabetes and hypertension. [SEIFA (Socio-Economic Indexes for Areas)]

**Table 2:** Associations between dentition status and incident cardiovascular disease among middle aged Australian women (n=8306)

|  | **HR<sup>a</sup>** | **95% CI<sup>b</sup>** | **p-value** |
| --- | --- | --- | --- |
| **Model 1** (unadjusted) | 0.70 | 0.62,0.78 | <0.001 |
| **Model 2** (adjusted for age, area of residence, SEIFA index of socio-economic disadvantage, SEIFA index of education, BMI, smoking status, diabetes and hypertension) | 0.83 | 0.74,0.93 | 0.001 |
| **Model 3** (adjusted for model 2 and diet quality) | 0.82 | 0.73,0.92 | <0.001 |

*SEIFA (Socio-Economic Indexes for Areas), BMI (Body Mass Index), <sup>a</sup>Hazarad Ratio, <sup>b</sup>Confidence interval.*

A functional dentition was associated with a higher diet quality [RR 1.12 (1.06, 1.20, p<0.001]. The relationship between dentition status and diet quality in those with a nonfunctional dentition was similar in women with dentures [RR 0.89 (0.83,0.95), p<0.001] and without dentures [RR 0.91 (0.78,1.05), p=0.22], (online supplemental tables 4 and 5).

Although not significant, high diet quality was related to a higher risk of incident CVD [ HR 1.11 (1.00, 1.24), p=0.06]. There was little evidence of association between the other measures of diet (total diet score as a continuous variable or diet score restricted to fruit and vegetable intake) and incident CVD [HR 1.00 (1.00, 1.01), p=0.63], [HR 1.09 (0.97, 1.21), p=0.14]), respectively (online supplemental table 6).

## Discussion

This study found that retention of teeth as part of a functional dentition was associated with a lower risk of future CVD. While retention of teeth was also associated with higher diet quality, adjusting for diet quality did not attenuate the relationship of teeth retention to future CVD. This finding was consistent when diet quality was restricted to fruit and vegetable intake. This suggests that diet quality is not the only pathway by which oral health, as measured by retention of teeth, is associated with CVD. It also specifically lends strength to the evidence that maintaining a functional dentition is an independent marker of better cardiovascular health, and that poor dentition is a marker of poor cardiovascular health among women.

The findings in this study that women with a non-functional dentition have a higher overall incidence of CVD (21.1% vs 15.5%) are consistent with the growing body of evidence that reports that the loss of teeth is associated with an increased risk of CVD,[7] and CVD related mortality.[7, 8, 23] Similarly, the dose response relationship reported in this study which showed that the risk of CVD decreased as the number of teeth increased is also consistent with the literature.[6, 7] An Australian study with a six-year follow-up reported that tooth loss was a risk marker for incident CVD related hospitalisation.[5] Very few studies however have analysed the impact of diet on the association between tooth loss and CVD. A study in male health professionals investigating the association between tooth loss and coronary heart disease adjusted for fibre and carrot intake reported a very slight attenuation in the relationship.[9] A longitudinal study in America [8] included diet quality defined by the Health Eating Index (HEI) as a covariate along with other established covariates in an analysis of the association between tooth loss and cardiovascular mortality and reported lower diet scores in those with ≥ 10 missing teeth.[8] Importantly, although studies consistently report that the loss of teeth increases the risk of CVD with a dose dependent relationship,[7, 8] and that the number of teeth retained is associated with a lower risk of CVD related mortality,[8, 23] they do not specifically report the association between the retention of a functional dentition (minimum of 20 teeth) on CVD risk.

In this study, women with a functional dentition were more likely to report a higher diet quality. These findings align with previous research indicating that the loss of a functional dentition impairs chewing function [24] and that tooth loss is associated with changes in food selection,[14] resulting in reduced intake of dietary protein and fibre as well as lower overall diet quality as measured by the HEI.[8] Interestingly, the presence of dentures did not change this relationship, indicating that dentures did not improve diet quality. This analysis is limited in that data collection did not include whether dentures restored the functional dentition (10 occluding pairs).

Diet quality has been shown to be associated with CVD,[15] and although not significant this study found an 11% increase in incident CVD for those with a high quality diet. This was unexpected, a previous study involving the same cohort using diet data from surveys 4-8 to investigate the relationship between diet quality and several non-communicable diseases including diabetes mellitus, coronary heart disease, hypertension, asthma, cancer, and depression failed to find an association between diet quality and coronary heart disease.[25] The sample size, the self-report bias of the diet survey, the fact that we did not specifically examine whether participants had a cardiovascular healthy diet and whether their diet changed over time may explain the association between diet and CVD reported in this study. Studies that have shown associations between diet and cardiovascular health have demonstrated the relationship of cardiovascular healthy diets such as the Mediterranean diet,[26] and the Dietary Approaches to Stop Hypertension (DASH) diet.[27]

Poor diet quality has been suggested as a key mechanism by which tooth loss leads to the increased risk of CVD.[11] However, this study’s findings indicate that dietary changes alone may not explain the impact of tooth loss on incident CVD. Systemic inflammation from oral diseases such as periodontitis may contribute to this association[10] and may also explain the other indicators of poor metabolic health such as higher rates of hypertension, increased BMI and diabetes noted in women with a non-functional dentition in this study.

## Strengths and Limitations

The study is a prospective study with 17 years of follow-up of a national sample of women, and although findings are not generalisable to men, there are fewer women represented in previous studies[7, 9, 28] and this study contributes to providing evidence for women. However, the women in the ALSWH tend to be healthier and of higher SES [18] therefore our study may underestimate the association between dentition status and incident CVD. Diet and tooth loss information were collected using surveys which may have resulted in self-reporting bias, additionally 13% of the dataset were excluded because of missingness which could have caused selection bias. Participants with missing diet and tooth data were older, more likely to live in a rural or remote location, more likely to have a low income and no formal education. However, there were no differences in the rates of diabetes, hypertension, incident CVD or fatal CVD. The analyses were unable to explore the role of systemic inflammation in the relation of dentition status and CVD, as inflammatory marker data was not collected, but this would be important to examine in future studies. Additional measures of diet quality such as the Mediterranean diet or DASH diet could not be analysed as survey 5 did not collect the necessary data to calculate these measures.

## Conclusions

In this cohort study of 8306 middle-aged women followed for up to 17 years the presence of a functional dentition was associated with a reduced risk of incident CVD and higher diet quality. The findings argue for both earlier interventions to maintain functional dentition and the recognition that the loss of a functional dentition may be a marker of poor future cardiovascular health and prompt cardiovascular prevention action.

## Acknowledgements

The research on which this paper is based was conducted as part of the Australian Longitudinal Study on Women’s Health by the University of Queensland and the University of Newcastle. We are grateful to the Australian Government Department of Health, Disability and Ageing for funding and to the women who provided the survey data.

The CCMS datasets are produced as part of the Australian Longitudinal Study on Women’s Health, by the University of Queensland and the University of Newcastle. We also acknowledge:

- The Australian Government Department of Health, Disability and Ageing for providing MBS, PBS and Aged Care data; and the Australian Institute of Health and Welfare (AIHW) as the integrating authority.
- The Data Linkage Unit at the Australian Institute of Health and Welfare (AIHW) for undertaking the data linkage to the National Death Index (NDI).
- The Centre for Health Record Linkage (CHeReL), NSW Ministry of Health and ACT Health, for the NSW Admitted Patients and Emergency Department Data Collections; and the ACT Admitted Patient Care and Emergency Department Data Collections.
- Queensland Health as the source for Queensland Hospital Admitted Patient and Emergency Data Collections; and the Statistical Analysis and Linkage Unit (Queensland Health) for the provision of data linkage.
- The Linkage, Data Outputs and Client Services teams at Data Linkage Services Western Australia as well as the Data Custodians of the WA Hospital Morbidity and Emergency Department Data Collections.
- SA NT DataLink, SA Health, and Northern Territory Department of Health, for the SA Public Hospital Separations, SA Public Hospital Emergency Department, NT Public Hospital Inpatient Activity and NT Public Hospital Emergency Department Data Collections.
- The Department of Health Tasmania, and the Tasmanian Data Linkage Unit, for the Public Hospital Admitted Patient Episodes and Tasmanian Emergency Department Presentations Data Collections.
- Victorian Department of Health as the source of the Victorian Admitted Episodes Dataset and the Victorian Emergency Minimum Dataset; and the Centre for Victorian Data Linkage (Victorian Department of Health) for the provision of data linkage.

The authors thank Professor Graham Giles and Professor Roger Milne of the Cancer Epidemiology Centre of Cancer Council Victoria, for permission to use the Dietary Questionnaire for Epidemiological Studies (Version 2), Melbourne: Cancer Council Victoria, 1996.

The authors also thank Ms Haeri Min for assisting with data programming.

## Data availability statement

ALSWH survey data are owned by the Australian Government Department of Health, Disability and Ageing and due to the personal nature of the data collected, release by ALSWH is subject to strict contractual and ethical restrictions. Ethical review of ALSWH is by the Human Research Ethics Committees at The University of Queensland and The University of Newcastle. De-identified data are available to collaborating researchers where a formal request to make use of the material has been approved by the ALSWH Data Access Committee. The committee is receptive of requests for datasets required to replicate results. Information on applying for ALSWH data is available from https://alswh.org.au/for-data-users/applying-for-data/. In addition, linked administrative data have been provided by third parties (see details here). In order for these linked data to be accessed through ALSWH, every data user must be added to the applicable Data Use Agreements and Human Research Ethics Committee protocols. Details of the HREC protocols covering the use of linked data are listed here.

## Author contributions

Dr Shalinie King: Contributed to conception, design, data acquisition and interpretation, drafted and critically revised the manuscript.

Dr Simone Marschner: Contributed to conception, design, statistical analysis, data interpretation and critically revised the manuscript.

Dr Desi Quintans: Contributed to programming, statistical analysis and critically revised the manuscript.

Dr Alice Gibson: Contributed to conception, manuscript drafting and critically revised the manuscript.

Dr Clara K Chow: Contributed to conception, design, data interpretation and critically revised the manuscript.

All authors gave final approval and agree to be accountable for all aspects of the work.

## Ethics statements

The ALSWH survey program has ongoing ethical approval from the Human Research Ethics Committees (HRECs) of the Universities of Newcastle and Queensland (approval numbers H076-0795, 2004/HE000224, respectively) for the 1946-51 cohort. The ALSWH also maintains institutional HREC approvals for external record linkage.

## References

1. World Health Organization. Cardiovascular diseases (CVD). [cited 2025 9th April 2025]; Available from: https://www.who.int/news-room/fact-sheets/detail/cardiovascular-diseases-(cvds).

2. Heart Foundation. Key statistics: Cardiovascular disease. [cited 2025 9th April 2025]; Available from: https://www.heartfoundation.org.au/your-heart/evidence-and-statistics/key-stats-cardiovascular-disease.

3. World Health Organization. Oral Health. [cited 2025 26th March 2025]; Available from: https://www.who.int/news-room/fact-sheets/detail/oral-health.

4. Do, L. and L. Luizzi, Tooth loss/Gum disease. In: ARCPOH. Australia’s Oral Health: National Study of Adult Oral Health 2017–18. 2019: Adelaide: The University of Adelaide, South Australia. p. 39–54/73-90.

5. Joshy, G., et al., Is poor oral health a risk marker for incident cardiovascular disease hospitalisation and all-cause mortality? Findings from 172 630 participants from the prospective 45 and Up Study. BMJ Open, 2016. 6(8): p. e012386.

6. Liljestrand, J.M., et al., Missing Teeth Predict Incident Cardiovascular Events, Diabetes, and Death. J Dent Res, 2015. 94(8): p. 1055–62.

7. Lee, H.J., et al., Tooth Loss Predicts Myocardial Infarction, Heart Failure, Stroke, and Death. Journal of Dental Research, 2019. 98(2): p. 164–170.

8. Shen, R., et al., Association between missing teeth number and all-cause and cardiovascular mortality: NHANES 1999–2004 and 2009–2014. Journal of Periodontology, 2024. 95(6): p. 571–581.

9. Joshipura, K.J., et al., Poor oral health and coronary heart disease. J Dent Res, 1996. 75(9): p. 1631–6.

10. Aoyama, N., et al., Associations among tooth loss, systemic inflammation and antibody titers to periodontal pathogens in Japanese patients with cardiovascular disease. Journal of Periodontal Research, 2018. 53(1): p. 117–122.

11. Moynihan, P.J., The relationship between nutrition and systemic and oral well-being in older people. The Journal of the American Dental Association, 2007. 138(4): p. 493–497.

12. Meyle, J. and I. Chapple, Molecular aspects of the pathogenesis of periodontitis. Periodontol 2000, 2015. 69(1): p. 7–17.

13. Yusuf, S., et al., Modifiable risk factors, cardiovascular disease, and mortality in 1551722 individuals from 21 high-income, middle-income, and low-income countries (PURE): a prospective cohort study. Lancet, 2020. 395(10226): p. 795–808.

14. Kossioni, A.E., The Association of Poor Oral Health Parameters with Malnutrition in Older Adults: A Review Considering the Potential Implications for Cognitive Impairment. Nutrients, 2018. 10(11).

15. Dong, C., et al., Cardiovascular disease burden attributable to dietary risk factors from 1990 to 2019: A systematic analysis of the Global Burden of Disease study. Nutrition, Metabolism and Cardiovascular Diseases, 2022. 32(4): p. 897–907.

16. Reynolds, A.N., et al., Dietary fibre in hypertension and cardiovascular disease management: systematic review and meta-analyses. BMC Medicine, 2022. 20(1): p. 139.

17. Qi, X.-X. and P. Shen, Associations of dietary protein intake with all-cause, cardiovascular disease, and cancer mortality: A systematic review and meta-analysis of cohort studies. Nutrition, Metabolism and Cardiovascular Diseases, 2020. 30(7): p. 1094–1105.

18. Dobson, A.J., et al., Cohort Profile Update: Australian Longitudinal Study on Women’s Health. Int J Epidemiol, 2015. 44(5): p. 1547,1547a-1547f.

19. Giles G and Ireland P, Dietary Questionnaire for Epidemiological Studies (Version 2) Melbourne. 1996: The Cancer Council Victoria; Melbourne, Australia.

20. Hodge, A., et al., The Anti Cancer Council of Victoria FFQ: relative validity of nutrient intakes compared with weighed food records in young to middle-aged women in a study of iron supplementation. Aust N Z J Public Health, 2000. 24(6): p. 576–83.

21. Collins, C.E., et al., The comparative validity and reproducibility of a diet quality index for adults: the Australian Recommended Food Score. Nutrients, 2015. 7(2): p. 785–98.

22. R Core Team 2024. R: A Language and Environment for Statistical Computing. R Foundation for Statistical Computing. Available from: https://www.R-project.org/].

23. Beukers, N., et al., Lower Number of Teeth Is Related to Higher Risks for ACVD and Death-Systematic Review and Meta-Analyses of Survival Data. Front Cardiovasc Med, 2021. 8: p. 621626.

24. Lahoud, T., A.Y.-D. Yu, and S. King, Masticatory Dysfunction in Older Adults: A Scoping Review. Journal of Oral Rehabilitation. n/a(n/a).

25. Hlaing-Hlaing, H., et al., Diet Quality and Incident Non-Communicable Disease in the 1946-1951 Cohort of the Australian Longitudinal Study on Women’s Health. Int J Environ Res Public Health, 2021. 18(21).

26. Martínez-González, M.A., A. Gea, and M. Ruiz-Canela, The Mediterranean Diet and Cardiovascular Health. Circulation Research, 2019. 124(5): p. 779–798.

27. Appel, L.J., et al., A clinical trial of the effects of dietary patterns on blood pressure. DASH Collaborative Research Group. N Engl J Med, 1997. 336(16): p. 1117–24.

28. Vedin, O., et al., Tooth loss is independently associated with poor outcomes in stable coronary heart disease. European Journal of Preventive Cardiology, 2020. 23(8): p. 839–846.

