## Supplementary material for "Tooth loss, diet and cardiovascular disease: A longitudinal study in middle-aged Australian women": online supplemental tables

**Online Supplement**

| **Table 1: Diet quality scores by dentition status** | | | |
| --- | --- | --- | --- |
| **Characteristic** | **Non-functional dentition** N = 2,558*^1^* | **Functional dentition** N = 5,748*^1^* | **p-value** |
| **ARFS score (0–74)** |  |  | **<0.001*^2^*** |
| Median (Q1, Q3) | 35 (29, 41) | 37 (31, 42) |  |
| Min – Max | 9 – 63 | 5 – 64 |  |
| Mean (SD) | 35 (9) | 37 (8) |  |
| **ARFS score (quintile)** |  |  | **<0.001*^3^*** |
| 1 | 650 (25%) | 1,012 (18%) |  |
| 2 | 510 (20%) | 1,151 (20%) |  |
| 3 | 488 (19%) | 1,173 (20%) |  |
| 4 | 454 (18%) | 1,207 (21%) |  |
| 5 | 456 (18%) | 1,205 (21%) |  |
| **ARFS score (binary)** |  |  | **<0.001*^3^*** |
| Low (Q1-Q3) | 1,648 (64%) | 3,336 (58%) |  |
| High (Q4-Q5) | 910 (36%) | 2,412 (42%) |  |
| **ARFS score (fruit/veg only; 0–36)** |  |  | **<0.001*^2^*** |
| Median (Q1, Q3) | 18 (13, 22) | 19 (14, 23) |  |
| Min – Max | 0 – 36 | 0 – 36 |  |
| Mean (SD) | 17 (7) | 18 (6) |  |
| **Fruit/veg score (quintile)** |  |  | **<0.001*^3^*** |
| 1 | 634 (25%) | 1,028 (18%) |  |
| 2 | 527 (21%) | 1,134 (20%) |  |
| 3 | 464 (18%) | 1,197 (21%) |  |
| 4 | 464 (18%) | 1,197 (21%) |  |
| 5 | 469 (18%) | 1,192 (21%) |  |
| **Fruit/veg score (binary)** |  |  | **<0.001*^3^*** |
| Low (Q1-Q3) | 1,625 (64%) | 3,359 (58%) |  |
| High (Q4-Q5) | 933 (36%) | 2,389 (42%) |  |
| *^1^*n (%) | | | |
| *^2^*Welch Two Sample t-test | | | |
| *^3^*Pearson's Chi-squared test  AFRS (Australian Recommended Food Score) | | | |

| **Table 2: Characteristics of the study population compared to women excluded due to missing diet or dentition data**. | | | |
| --- | --- | --- | --- |
| **Characteristic** | **Eligible** N = 8,306^1^ | **Excluded**  N = 850*^1^* | **p-value***^2^* |
| **Age** | 58.5 (1.5) | 58.6 (1.5) | **0.001** |
| **Area of residence** |  |  | **<0.001** |
| Major cities | 3,186 / 8,302 (38.4%) | 253 / 850 (29.8%) |  |
| Inner regional | 3,221 / 8,302 (38.8%) | 366 / 850 (43.1%) |  |
| Outer regional | 1,594 / 8,302 (19.2%) | 196 / 850 (23.1%) |  |
| Remote/Very remote/Overseas | 301 / 8,302 (3.6%) | 35 / 850 (4.1%) |  |
| (Missing) | 4 | 0 |  |
| **Household income** |  |  | **<0.001** |
| $0-$25999 annually | 1,092 / 6,604 (16.5%) | 130 / 614 (21.2%) |  |
| $26000-$51999 annually | 1,990 / 6,604 (30.1%) | 202 / 614 (32.9%) |  |
| >= $52,000 annually | 2,352 / 6,604 (35.6%) | 155 / 614 (25.2%) |  |
| Don't know | 249 / 6,604 (3.8%) | 34 / 614 (5.5%) |  |
| Don't want to answer | 492 / 6,604 (7.5%) | 54 / 614 (8.8%) |  |
| Income same as mine | 429 / 6,604 (6.5%) | 39 / 614 (6.4%) |  |
| (Missing) | 1,702 | 236 |  |
| **Education level** |  |  | **<0.001** |
| No qualification | 1,196 / 8,249 (14.5%) | 222 / 834 (26.6%) |  |
| High school/Trade | 4,282 / 8,249 (51.9%) | 449 / 834 (53.8%) |  |
| Certificate/Diploma | 1,424 / 8,249 (17.3%) | 99 / 834 (11.9%) |  |
| University | 1,347 / 8,249 (16.3%) | 64 / 834 (7.7%) |  |
| (Missing) | 57 | 16 |  |
| **SEIFA index of socioeconomic disadvantage** | 4,258 / 8,296 (51.3%) | 356 / 850 (41.9%) | **<0.001** |
| (Missing) | 10 | 0 |  |
| **SEIFA index of education** | 4,245 / 8,296 (51.2%) | 366 / 850 (43.1%) | **<0.001** |
| (Missing) | 10 | 0 |  |
| **BMI** | 27.2 (5.4) | 27.7 (5.8) | 0.12 |
| (Missing) | 79 | 22 |  |
| **Diabetes** | 451 / 8,297 (5.4%) | 59 / 847 (7.0%) | 0.07 |
| (Missing) | 9 | 3 |  |
| **Hypertension** | 2,146 / 8,297 (25.9%) | 240 / 847 (28.3%) | 0.12 |
| Unknown | 9 | 3 |  |
| **Smoking status** |  |  | **0.023** |
| Never | 5,052 / 8,300 (60.9%) | 511 / 841 (60.8%) |  |
| Ex-smoker | 2,394 / 8,300 (28.8%) | 220 / 841 (26.2%) |  |
| Current | 854 / 8,300 (10.3%) | 110 / 841 (13.1%) |  |
| Unknown | 6 | 9 |  |
| **All-cause mortality** | 513 / 8,306 (6.2%) | 62 / 850 (7.3%) | 0.2 |
| **Incident CVD** | 1,432 / 8,306 (17.2%) | 160 / 850 (18.8%) | 0.2 |
| - Fatal CVD | 36 / 1,432^#^ (2.5%) | 7 / 160^#^ (4.4%) | 0.2 |
| *^1^* n / N (%); Mean (SD) | | | |
| *^2^* Pearson’s Chi-squared test; Welch Two Sample t-test | | | |
| *^#^ Participants with incident CVD* | | | |

| **Table 3: Association between dentition status and cardiovascular disease** | | | |
| --- | --- | --- | --- |
| **Model 2** |  |  |  |
|  | *HR^1^* | *CI^2^* | ***P*** |
| **Dentition status** |  |  |  |
| Non-functional dentition | - | - | - |
| Functional dentition | 0.83 | 0.74, 0.93 | **0.001** |
| **Age** | 1.09 | 1.05, 1.13 | **< 0.001** |
| **Area of residence** |  |  |  |
| Inner Regional | - | - | - |
| Remote/very remote/overseas | 0.767 | 0.519,1.133 | 0.183 |
| Urban Cities | 0.924 | 0.798, 1.070 | 0.292 |
| Outer Regional | 0.880 | 0.739, 1.047 | 0.150 |
| **SEIFA index of socioeconomic disadvantage** |  |  |  |
| Low socio-economic disadvantage | - | - | - |
| High socio-economic disadvantage | 1.00 | 0.87, 1.15 | 0.98 |
| **SEIFA index of education** |  |  |  |
| Low index of education and occupation | - | - | - |
| High index of education and occupation | 0.90 | 0.77, 1.03 | 0.11 |
| **BMI** | 1.02 | 1.01, 1.03 | **0.003** |
| **Smoking status** |  |  |  |
| Never -smoked | - | - | - |
| Ex -smoker | 1.16 | 1.03, 1.31 | 0.014 |
| Current smoker | 1.65 | 1.40, 1.95 | **<0.001** |
| **Diabetes** |  |  |  |
| No | - | - | - |
| Yes | 1.56 | 1.28, 1.89 | **<0.001** |
| **Hypertension** |  |  |  |
| No | - | - | - |
| Yes | 1.40 | 1.24, 1.58 | **<0.001** |
| **Model 3** |  |  |  |
|  | *HR^1^* | *CI^2^* | ***P*** |
| **Dentition status** |  |  |  |
| Non-functional dentition | - | - | - |
| Functional dentition | 0.82 | 0.73, 0.92 | **<0.001** |
| **Diet (bottom three quintiles – low quality)** | - | - | - |
| **Diet (top two quintiles- high quality)** | 1.12 | 1.01, 1.25 | **0.004** |
| **Age** | 1.09 | 1.05, 1.13 | < 0.001 |
| **Area of residence** |  |  |  |
| Inner Regional | - | - | - |
| Remote/very remote/overseas | 0.767 | 0.519, 1.133 | 0.182 |
| Urban Cities | 0.926 | 0.799, 1.073 | 0.304 |
| Outer Regional | 0.882 | 0.740, 1.050 | 0.157 |
| **SEIFA index of socioeconomic disadvantage** |  |  |  |
| Low socio-economic disadvantage | - | - | - |
| High socio-economic disadvantage | 1.00 | 0.87, 1.16 | >0.9 |
| **SEIFA index of education** |  |  |  |
| Low index of education and occupation | - | - | - |
| High index of education and occupation | 0.89 | 0.77, 1.02 | 0.092 |
| **BMI** | 1.01 | 1.00, 1.03 | **0.003** |
| **Smoking status** |  |  |  |
| Never -smoked | - | - | - |
| Ex -smoker | 1.16 | 1.03, 1.31 | **0.014** |
| Current smoker | 1.68 | 1.42, 1.98 | **<0.001** |
| **Diabetes** |  |  |  |
| No | - | - | - |
| Yes | 1.55 | 1.27, 1.88 | **<0.001** |
| **Hypertension** |  |  |  |
| No | - | - | - |
| Yes | 1.40 | 1.24, 1.58 | **<0.001** |
| *^1^*HR = Hazard Ratio, ^2^CI = Confidence Interval | | | |

| **Table 4: Association between dentition status and diet quality** |  |  |  |
| --- | --- | --- | --- |
| **Characteristic** | **RR** | **95% CI** | **p-value** |
| **Dentition group** |  |  |  |
| Non-functional dentition | — | — |  |
| Functional dentition | 1.12 | 1.06, 1.20 | **<0.001** |
| **Area of residence** |  |  |  |
| Inner regional | — | — |  |
| Remote/Very remote/Overseas | 0.92 | 0.76, 1.09 | 0.392 |
| Major cities | 0.98 | 0.91, 1.05 | 0.509 |
| Outer regional | 0.95 | 0.87, 1.04 | 0.286 |
| **age** | 1.02 | 1.01, 1.04 | **0.007** |
| **SEIFA index of socioeconomic disadvantage** |  |  |  |
| Low socio-economic disadvantage | — | — |  |
| High socio-economic disadvantage | 1.01 | 0.94, 1.08 | 0.782 |
| **SEIFA index of education** |  |  |  |
| Low index of education and occupation | — | — |  |
| High index of education and occupation | 1.05 | 0.98, 1.13 | 0.134 |
| **BMI** | 1.00 | 0.99, 1.00 | 0.132 |
| **Smoking status** |  |  |  |
| Never | — | — |  |
| Ex-smoker | 0.98 | 0.92, 1.03 | 0.404 |
| Current | 0.68 | 0.60, 0.76 | **<0.001** |
| **Diabetes** |  |  |  |
| No | — | — |  |
| Yes | 1.12 | 0.99, 1.25 | 0.061 |
| **Hypertension** |  |  |  |
| No | — | — |  |
| Yes | 1.00 | 0.94, 1.06 | 0.990 |
| Abbreviations: CI = Confidence Interval, RR = Relative Risk | | | |

| **Table 5: Association between dentition status and diet for women with and without dentures** | | | |
| --- | --- | --- | --- |
| **Characteristic** | **RR***^1^* | **95% CI***^1^* | **p-value** |
| **Dentition status** |  |  |  |
| **Functional dentition** | — | — |  |
| Non-functional dentition with dentures | 0.89 | 0.83,0.95 | <0.001 |
| Non-functional dentition without dentures | 0.91 | 0.78, 1.05 | 0.22 |
| **Area of residence** |  |  |  |
| Inner regional | — | — | — |
| Remote/Very remote/Overseas | 0.92 | 0.76, 1.09 | 0.392 |
| Outer regional | 0.95 | 0.87, 1.04 | 0.286 |
| Remote | 1.06 | 0.89, 1.23 | 0.5 |
| Major cities | 0.98 | 0.91, 1.05 | 0.509 |
| **Age** | 1.03 | 1.01, 1.04 | **0.007** |
| **SEIFA index of socioeconomic disadvantage** | 1.02 | 1.00, 1.04 | 0.04 |
| Low socio-economic disadvantage | — | — |  |
| High socio-economic disadvantage | 1.01 | 0.94, 1.08 | 0.78 |
| **SEIFA index of education** |  |  |  |
| Low index of education and occupation | — | — |  |
| High index of education and occupation | 1.05 | 0.98, 1.13 | 0.14 |
| **BMI** | 1.00 | 0.99, 1.00 | 0.13 |
| **Smoking status** |  |  |  |
| Never | — | — |  |
| Ex-smoker | 0.98 | 0.92, 1.03 | 0.40 |
| Current | 0.68 | 0.60, 0.76 | **<0.001** |
| **Diabetes** |  |  |  |
| No | — | — |  |
| Yes | 1.12 | 0.99, 1.25 | 0.06 |
| **Hypertension** |  |  |  |
| No | — | — |  |
| Yes | 1.00 | 0.94, 1.07 | > 0.9 |

Abbreviations: CI = Confidence Interval, RR = Relative Risk

| **Table 6: Association between diet quality and cardiovascular disease** | | | |
| --- | --- | --- | --- |
| **Characteristic** | **HR***^1^* | **95% CI***^1^* | **p-value** |
| **Diet quality (Binary)** |  |  |  |
| **Diet quality** | 1.11 | 1.00, 1.24 | 0.051 |
| **Area of residence** |  |  |  |
| Inner regional | — | — |  |
| Remote/Very remote/Overseas | 0.76 | 0.52, 1.13 | 0.176 |
| Outer regional | 0.88 | 0.74, 1.05 | 0.168 |
| Major cities | 0.91 | 0.79, 1.06 | 0.227 |
| **Age** | 1.09 | 1.05, 1.13 | **<0.001** |
| **SEIFA index of socioeconomic disadvantage** |  |  |  |
| Low socio-economic disadvantage | — | — |  |
| High socio-economic disadvantage | 1.00 | 0.87, 1.15 | 0.984 |
| **SEIFA index of education** |  |  |  |
| Low index of education and occupation | — | — |  |
| High index of education and occupation | 0.87 | 0.76, 1.01 | 0.062 |
| **BMI** | 1.02 | 1.01, 1.03 | **0.001** |
| **Smoking status** |  |  |  |
| Never | — | — |  |
| Ex-smoker | 1.17 | 1.04, 1.32 | **0.011** |
| Current | 1.72 | 1.46, 2.02 | **<0.001** |
| **Diabetes** |  |  |  |
| No | — | — |  |
| Yes | 1.57 | 1.29, 1.91 | **<0.001** |
| **Hypertension** |  |  |  |
| No | — | — |  |
| Yes | 1.40 | 1.24, 1.58 | **<0.001** |
| **Diet quality (continuous)** |  |  |  |
| **Diet quality** | 1.00 | 1.00, 1.01 | 0.63 |
| **Area of residence** |  |  |  |
| Inner regional | — | — | — |
| Remote/Very remote/Overseas | 0.50 | 1.7, 0.68 | 0.405 |
| Outer regional | 0.49 | 1.1, 0.09 | 0.101 |
| Major cities | 0.48 | 0.97, 0.01 | 0.054 |
| **Age** | 1.09 | 1.05, 1.13 | **<0.001** |
| **SEIFA index of socioeconomic disadvantage** |  |  |  |
| Low socio-economic disadvantage | — | — |  |
| High socio-economic disadvantage | 1.00 | 0.87, 1.15 | > 0.9 |
| **SEIFA index of education** |  |  |  |
| Low index of education and occupation | — | — |  |
| High index of education and occupation | 0.87 | 0.76, 1.00 | 0.05 |
| **BMI** | 1.02 | 1.01, 1.03 | **<0.001** |
| **Smoking status** |  |  |  |
| Never | — | — |  |
| Ex-smoker | 1.18 | 1.04, 1.33 | **0.008** |
| Current | 1.75 | 1.49, 2.06 | **<0.001** |
| **Diabetes** |  |  |  |
| No | — | — |  |
| Yes | 1.58 | 1.30, 1.92 | **<0.001** |
| **Hypertension** |  |  |  |
| No | — | — |  |
| Yes | 1.40 | 1.24, 1.58 | **<0.001** |
| **Fruit and vegetable intake only** |  |  |  |
| **Diet quality** | 1.09 | 0.97, 1.21 | 0.141 |
| **Area of residence** |  |  |  |
| Inner regional | — | — |  |
| Remote/Very remote/Overseas | 0.76 | 0.52, 1.13 | 0.174 |
| Outer regional | 0.88 | 0.74, 1.05 | 0.162 |
| Major cities | 0.91 | 0.79, 1.06 | 0.229 |
| **Age** | 1.09 | 1.05, 1.13 | **<0.001** |
| **SEIFA index of socioeconomic disadvantage** |  |  |  |
| Low socio-economic disadvantage | — | — |  |
| High socio-economic disadvantage | 1.00 | 0.87, 1.15 | 0.970 |
| **SEIFA index of education** |  |  |  |
| Low index of education and occupation | — | — |  |
| High index of education and occupation | 0.87 | 0.75, 1.00 | **0.050** |
| **BMI** | 1.02 | 1.01, 1.03 | **<0.001** |
| **Smoking status** |  |  |  |
| Never | — | — |  |
| Ex-smoker | 1.17 | 1.04, 1.32 | **0.009** |
| Current | 1.76 | 1.49, 2.06 | **<0.001** |
| **Diabetes** |  |  |  |
| No |  |  |  |
| Yes | 1.58 | 1.30, 1.93 | **<0.001** |
| **Hypertension** |  |  |  |
| No |  |  |  |
| Yes | 1.40 | 1.24, 1.58 | **<0.001** |

*^1^*HR = Relative Risk, CI = Confidence Interval
